# Comparison of respiratory protection during exercising tasks between different wearing methods of replaceable particulate respirators and powered air-purifying respirators

**DOI:** 10.1101/2021.05.15.21257205

**Authors:** Hiroka Baba, Hajime Ando, Kazunori Ikegami, Shingo Sekoguchi, Taiki Shirasaka, Akira Ogami

**Author notes:** Corresponding author: Hajime Ando, MD, PhD., Department of Work System and Health at the University of Occupational and Environmental Health, Japan1-1, Iseigaoka, Yahata-nishi-ku, Kitakyushu, Fukuoka 807-8555, Japan.

## Abstract

**Background:** This study evaluated the differences in respiratory protection between replaceable particulate respirators (RPRs) and powered air-purifying respirators (PAPRs), with different wearing methods, during exercising tasks.

**Methods:** Ten participants wore either RPR or PAPR according to the recommended method, with a knit cover placed between the facepiece cushion and the face, or with the headband on a helmet. We measured the number of particles inside and outside the respiratory protective equipment (RPE) during exercising tasks for each wearing variation. The exercise state was set to exercise with an ergometer set at 80W load. While exercising tasks, the participants performed five actions adopted from JIS T8150 in 2018 (1. Normal breathing, 2. Deep breathing, 3. Turning head side to side, 4. Moving head up and down, 5. Talking). Each action was performed for 1 min. For measurements of exercise state, after 10 minutes of exercise tasks, we measured while the exercise was continued. The fit factor was calculated by dividing the concentration within the RPE by the concentration outside of it. Data were analyzed after they were log-transformed with a linear mixed model, with fit factor as the dependent variable.

**Results:** We compared the results with experimental data of resting state reported in our previous studies. Fit factor of RPRs in the exercise state was significantly lower (p<0.001) than that in the resting state, indicating inadequate respiratory protection. In contrast, the fit factor of PAPRs during exercising tasks was significantly lower (p<0.001) than that at rest; however, respiratory protection was maintained. PAPR did not show a significant decrease (p=1.000) in fit factor owing to the wearing variations during exercising tasks.

**Conclusions:** PAPRs were found to be superior to RPRs in terms of respiratory protection. PAPRs are better than RPRs for workers who have to wear RPE inappropriately due to health problems.

## Introduction

The number of cases of pneumoconiosis and occupational diseases due to complications of pneumoconiosis in Japan decreased to 164 in 2019; however, new cases are still occurring.^1^

It is necessary to implement technical and work control measures to reduce the risk of pneumoconiosis and its complications. If such measures are not sufficient to reduce hazards, it is important to wear respiratory protective equipment (RPE), which helps to reduce workers’ exposure to dust and toxic chemicals through inhalation and prevent various occupational diseases. However, previous studies have shown that wearing RPE that do not allow sufficient adhesion between the face and the RPE does not provide adequate respiratory protection.^2,3^ For this reason, the Ministry of Health, Labour and Welfare in Japan has issued guidelines on matters to be considered in the selection and use of RPE.^4^ Previous studies have shown that those working in dusty workplaces in Japan wear RPE in various ways. We found that RPE was worn in several inappropriate ways, such as by wearing a tightening strap over the helmet or by wearing RPE over a towel wrapped around the face in hot workplace environments.^5^ When the respiratory protection capacity of RPE is reduced, inhalation of dust, toxic chemicals, etc. can lead to the development of respiratory diseases such as pneumoconiosis and lung cancer. Prior to this study, we evaluated the difference in protective performance between replaceable particulate respirators (RPRs) and powered air-purifying respirators (PAPRs) by having the participants wear them at rest in various ways in a laboratory. In this previous study, the average leakage rate at rest for RPR ranged from 1.82% to 10.92%, and the average leakage rate at rest for PAPR ranged from 0.18% to 0.42%. Under the rest condition, while there was a decrease in respiratory protection with RPR due to different methods of wear, no significant decrease in respiratory protection was observed with PAPR based on different methods of wear. This indicates that PAPRs can provide sufficient protection for workers who are unable to wear RPRs appropriately due to the work environment.^6^ A previous study that examined the effect of the wearing method on respiratory protection at rest found that PAPRs maintained high respiratory protection regardless of the wearing method; therefore, we thought it might be possible to maintain high respiratory protection regardless of workload. In this previous study, we were unable to verify the effect of workload on respiratory protection. Therefore, we thought it was important to verify the effect of RPE on respiratory protection during workload, depending on the method of wearing RPE among workers. The purpose of this study was to measure the fit factor of RPRs and PAPRs during workload, in a laboratory, using various wearing methods, and to clarify the differences in respiratory protection and the wearing methods that maintain adequate respiratory protection.

## Methods

### Ethical Approval

The Ethics and Informed Consent Procedure for this study was approved by the Ethics Committee of Medical Research, University of Occupational and Environmental Health, Japan (Receipt No. H30-58). Informed consent was obtained from all the participants.

### Study design and setting

We conducted a crossover comparison study of 10 participants who agreed to participate in the experiment. Two physical states (resting and exercise state) were used to measure the fit factor of each combination (hereafter called “wearing variation”) of the RPE type and wearing method.

Fit factor is the numerical result of a quantitative fit test performed on an RPE face piece, which indicates the effectiveness of the seal against the face.

The experiment was conducted from August to September 2018 in an artificial climate chamber at the University of Occupational and Environmental Health, Japan to exclude environmental influences. The climatic conditions in the artificial climate chamber were set to maintain a room temperature of 20°C and a relative humidity of 50%.

### Participants

We recruited participants from the University of Occupational and Environmental Health, Japan. All participants were healthy adults over 20 years old and were non-smokers, to prevent tobacco dust from affecting the leakage rate measurement. Ten participants, eight males (mean age [standard deviation (SD)]: 32.1 [3.98] years) and two females (mean age [SD]: 34.0 [5.0] years), were eligible for this study.

### Particulate respirator

The RPR selected for testing was the 1180–05 (Koken Ltd.), and the PAPR was the BL–321S (Koken Ltd). The RPR used was a model that complied with RL2, and the particulate filtering efficiency according to the Japanese standard for dust mask is 95%.^7^

PAPR used was a model that complied with PL1, and the particulate filtering efficiency in the Japanese standard for PAPRs is 95%.^8^ The PAPR used had the following capacity: motor blower capacity; large airflow volume type (over 138 L/min), leakage rate; B class (less than 5.0%), filtering efficiency; PL1 (over 95.0%). The RPR selected for testing was the 1180–05 (Koken Ltd., Japan) with a filtering efficiency RL2 (over 95.0%). This RPR consisted of a half facepiece and a single filter with a shape similar to that of the BL–321S.

To minimize the influence of individual RPE fitting techniques, the participants used a mirror to standardize the fitting procedure. The tightness of the fastening straps was measured with a measuring instrument using a Sensor Interchangeable Amplifier (force gauge) (eZT, IMADA CO., LTD, Japan). The tightness of the fastening straps was adjusted so that the force with which the fastening straps of the RPE pressurized the head was uniform.

### Wearing variations

The method of wearing the RPEs was based on the commonly observed method of wearing them in the workplace^5^ revealed in our previous study. They were as follows: the recommended method (R) in which the headband was placed on the area from the parietal region to the occipital region with nothing between the facepiece cushion and the face; the method in which a knit cover is placed between the facepiece cushion and the face (K); and the method in which the headband was placed over a helmet (H). Figure 1 shows an image of the wearing methods.

**Figure 1.**
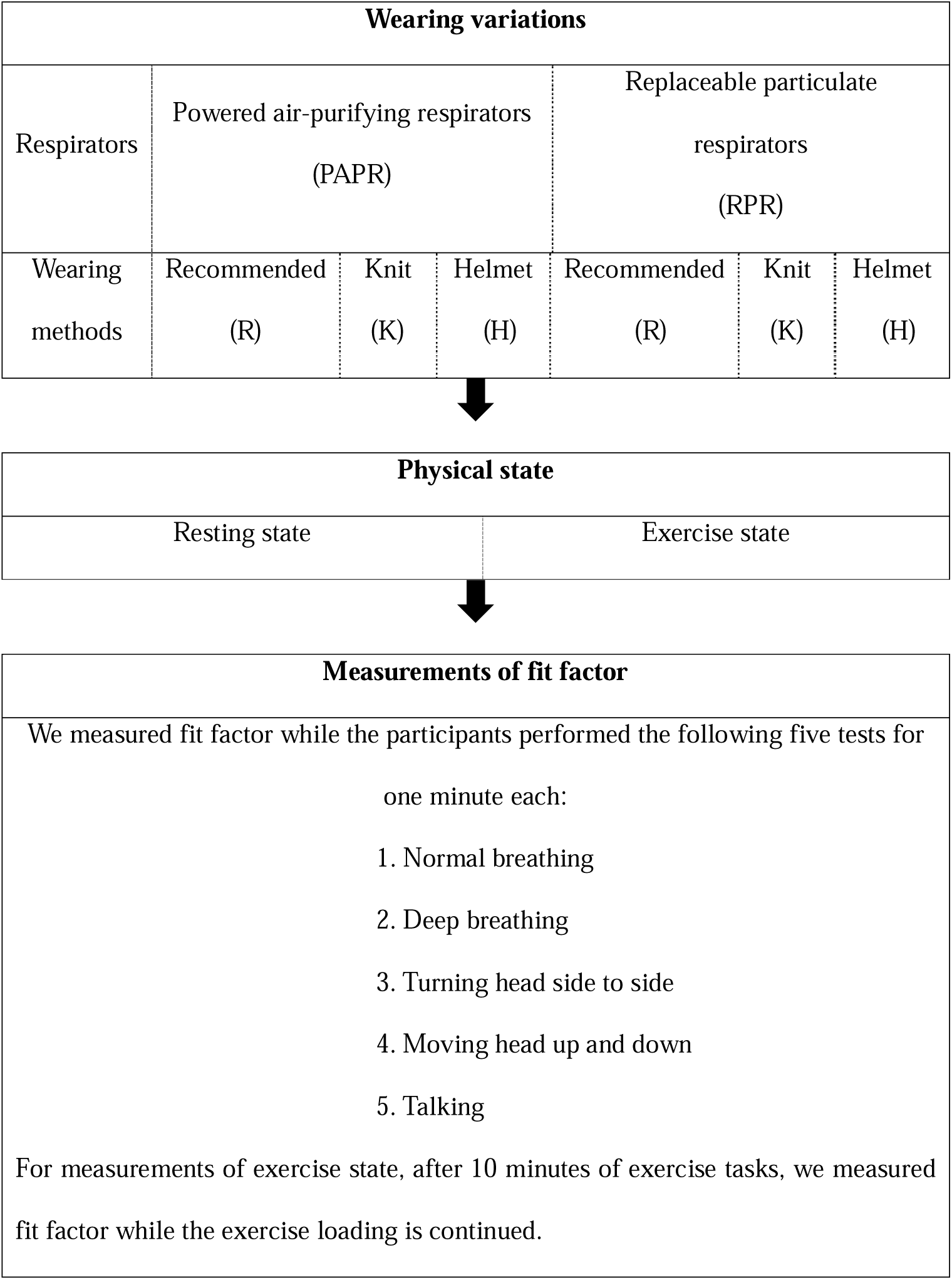
Photographs depicting different wearing methods, using a replaceable particulate respirator as an example. We adapted these photos from the study by Sekoguchi et al.^6^

We represented wearing variation as the combination of “RPR” or “PAPR” and “wearing method.” For example, a PAPR worn according to the recommended method (R) was represented with PAPR-R. We conducted an experiment with six wearing variations. Finally, we set RPR-R, RPR-K, RPR-H, PAPR-R, PAPR-K, and PAPR-H as the six wearing variations.

### Measurement procedure

We measured the fit factor of the RPE at two physical states, resting and exercise, for the six different wearing variations. We reported the experimental data of the resting state in a previous study.^6^ The exercise state was set to exercise with an ergometer set at an 80 W load. The order of the fit factor measurements for each wearing variation was assigned using a random number table. Figure 2 illustrates the measurement procedure.

**Figure 2.** Outline of the study

### Measurement of fit factor

To measure fit factor, the participants wore the particulate respirator and performed five actions (1. Normal breathing, 2. Deep breathing, 3. Turning the head side-to-side: 4. Moving head up and down, and 5. Talking). Each action was performed for 1 min. For measurements of exercise state, after 10 minutes of exercise tasks, we measured fit factor while the exercise loading is continued. The measurement device measured the concentration of atmospheric dust inside and outside the particulate respirator.

The fit testing procedure according to the occupational safety and health administration (OSHA) specifies that seven motions should be performed to measure the fit factor.^9^ However, because the measurement was to be performed while exercising tasks, the five motions of the Japan Industrial Standard (JIS) T8150 in 2018 were adopted for safety reasons.^10^ To avoid any influence on the measurements due to the fatigue in the participants, fit factor measurements while exercising tasks were limited to two times per day, with an interval of at least one hour.

### Measurement apparatus of fit factor

The leakage rate was measured using an MT–03 device (Sibata Scientific Technology Ltd.), which uses a light scattering system particle counter for the detection part and counts the particles present in the air inside and outside the RPE sucked at 1 l/min. After measuring the air outside the RPE for 17 s, the instrument measured the air inside the RPE for 17 s. The replacement time for the dust remaining in the pipe at the start of the measurement and when switching the measurement path was set to 10 s. The time required for each measurement was approximately 1 min. The particles measured by the device were atmospheric dust with a particle size of at least 0.5 μm. During the measurement, incense sticks were burned to maintain a level of dust in an environment of more than 1000 counts/3 s, which is the recommended value for MT-03. The concentration within the RPE was measured by sampling the air inside the facepiece using a tube joint set fixed to the sampling tube and the RPE. The concentration outside the RPE was measured by sampling the air outside the RPE using a sampling tube fixed with a string hung from the ceiling so that the end of the sampling tube was close to the RPE.

### Calculating the fit factor

According to JIS T8150^10^ or fit testing procedure by OSHA,^9^ the fit factor was calculated by dividing the concentration within the particulate respirator (Ni) by the concentration outside it (No):

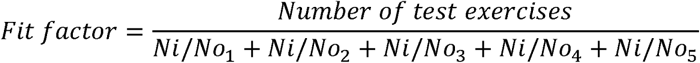

The numbers from 1 to 5 represent test exercises 1-5.

OSHA indicated that the test participant should not be permitted to wear a half mask or quarter facepiece respirator unless a minimum fit factor of 100 is obtained.^9^ In this study, we used fit factor ≥ 100 as the criterion to ensure that respiratory protection was maintained.

### Variables

The outcome variable was fit factor, and the predictor variables were physical state (resting state and exercise state) and wearing variations. We adjusted for sex as a confounding factor.

### Statistical methods

Data were analyzed after they were log-transformed with a linear mixed model (LMM), with fit factor as the dependent variable. Among the independent variables, the random factor was the survey participants, and the fixed factors were sex, physical states, wearing variations, and interaction between physical states and wearing variations. The Bonferroni method was used for multiple comparisons. The estimated marginal means (EMM) by physical states or wearing variations were calculated by adjusting for the dependent variable of LMM. All statistical analyses were performed using the IBM SPSS Statistics 23.0. The significance level was set at p<0.05.

## Results

### Fit factor according to physical states and wearing variations

Table 1 shows the number of cases with fit factor > 100 and the mean (SD) of fit factor for each physical condition and wearing variation.

**Table 1.**
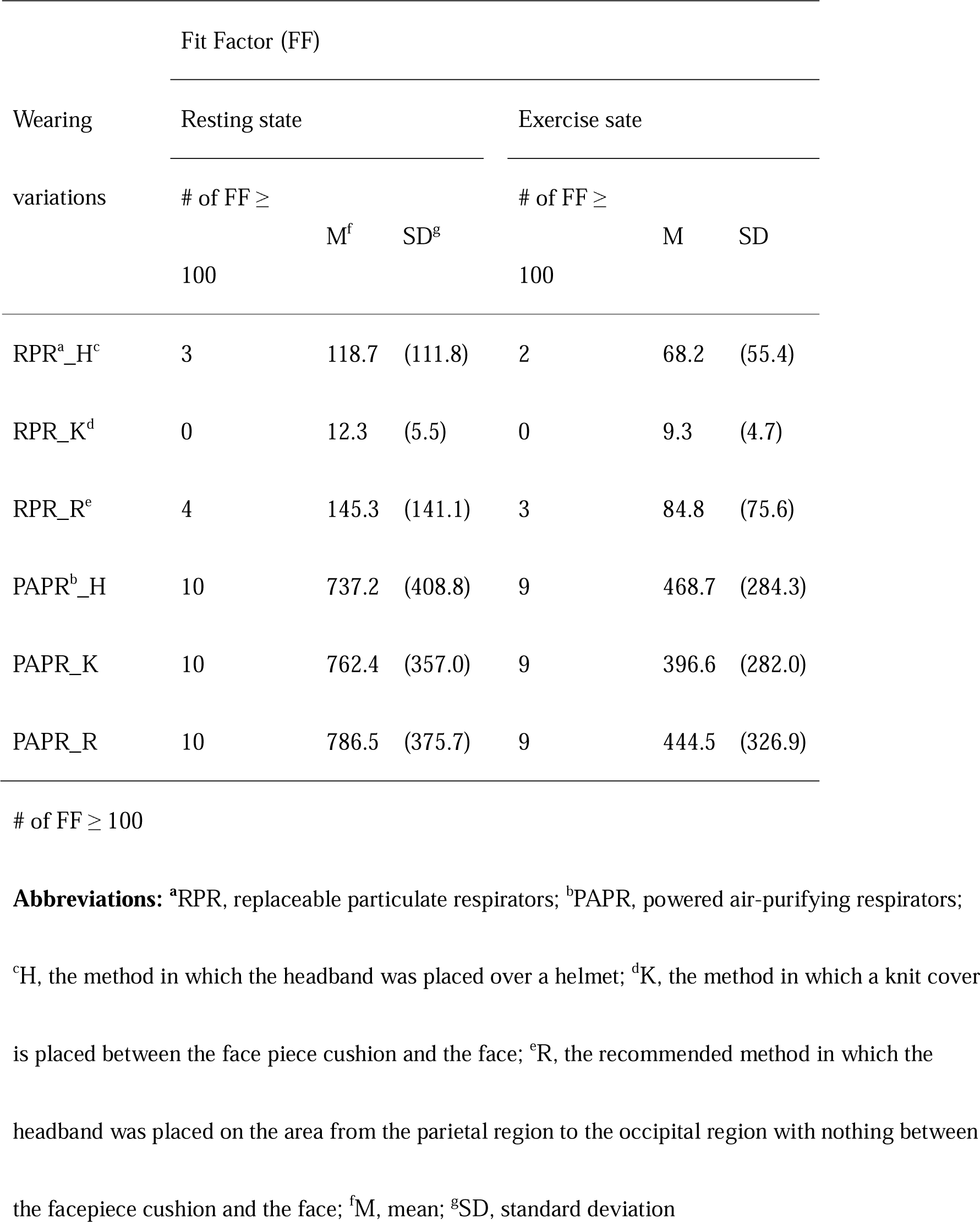
Number of cases with fit factor > 100 and the mean (standard deviation) of fit factor for each physical condition and wearing variation.

| Wearing variations | Fit Factor (FF) |  |  |  |  |  |
| --- | --- | --- | --- | --- | --- | --- |
|  | Resting state |  |  | Exercise state |  |  |
|  | # of FF ≥ | M<sup>f</sup> | SD<sup>g</sup> | # of FF ≥ | M | SD |
|  | 100 |  |  | 100 |  |  |
| RPR<sup>a</sup>_H<sup>c</sup> | 3 | 118.7 | (111.8) | 2 | 68.2 | (55.4) |
| RPR_K<sup>d</sup> | 0 | 12.3 | (5.5) | 0 | 9.3 | (4.7) |
| RPR_R<sup>e</sup> | 4 | 145.3 | (141.1) | 3 | 84.8 | (75.6) |
| PAPR<sup>b</sup>_H | 10 | 737.2 | (408.8) | 9 | 468.7 | (284.3) |
| PAPR_K | 10 | 762.4 | (357.0) | 9 | 396.6 | (282.0) |
| PAPR_R | 10 | 786.5 | (375.7) | 9 | 444.5 | (326.9) |

We could not observe cases of fit factor ≥ 100 when the participants wore the RPE by RPR_K. The number of cases with fit factor ≥ 100 was higher when wearing the PAPR than when wearing the RPR in both the resting state and exercise state. In each of the three methods, when wearing the PAPR in the resting state, the fit factor was ≥100; however, only one of ten cases had fit factor <100 when wearing the PAPR in exercise state. The mean values of fit factor were also higher when wearing the PAPR than when wearing the RPR, and they were higher in the resting state than in the exercise state.

### Comparisons of the values of fit factor by physical states and wearing variations

Table 2 shows the results of the statistical comparison of the values of fit factor by physical states and wearing variations. We analyzed the interactions between the physical states and wearing variations.

**Table 2.**
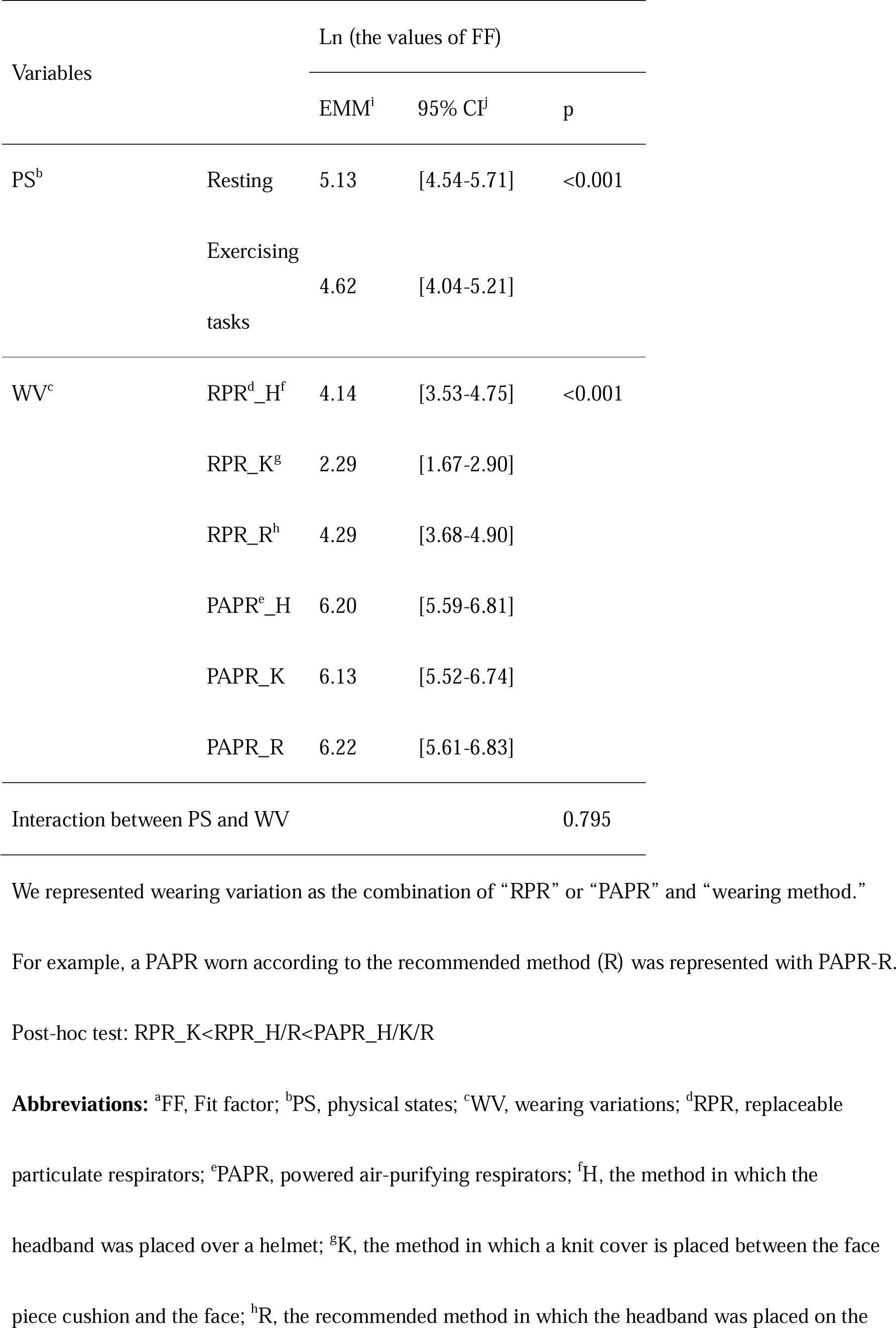
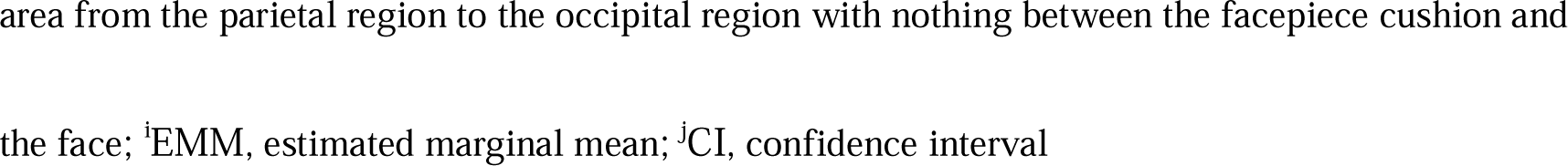
Comparison of the values of FF^a^ among physical states and among wearing variations

| Variables |  | Ln (the values of FF) |  |  |
| --- | --- | --- | --- | --- |
|  |  | EMM<sup>i</sup> | 95% CI<sup>j</sup> | p |
| PS<sup>b</sup> | Resting | 5.13 | [4.54-5.71] | <0.001 |
|  | Exercising tasks | 4.62 | [4.04-5.21] |  |
| WV<sup>c</sup> | RPR<sup>d</sup>_H<sup>f</sup> | 4.14 | [3.53-4.75] | <0.001 |
|  | RPR_K<sup>g</sup> | 2.29 | [1.67-2.90] |  |
|  | RPR_R<sup>h</sup> | 4.29 | [3.68-4.90] |  |
|  | PAPR<sup>e</sup>_H | 6.20 | [5.59-6.81] |  |
|  | PAPR_K | 6.13 | [5.52-6.74] |  |
|  | PAPR_R | 6.22 | [5.61-6.83] |  |
| Interaction between PS and WV |  |  |  | 0.795 |

We represented wearing variation as the combination of “RPR” or “PAPR” and “wearing method.”
For example, a PAPR worn according to the recommended method (R) was represented with PAPR-R.
Post-hoc test: RPR\_K<RPR\_H/R<PAPR\_H/K/R
**Abbreviations:** <sup>a</sup>FF, Fit factor; <sup>b</sup>PS, physical states; <sup>c</sup>WV, wearing variations; <sup>d</sup>RPR, replaceable
particulate respirators; <sup>e</sup>PAPR, powered air-purifying respirators; <sup>f</sup>H, the method in which the
headband was placed over a helmet; <sup>g</sup>K, the method in which a knit cover is placed between the face
piece cushion and the face; <sup>h</sup>R, the recommended method in which the headband was placed on the
area from the parietal region to the occipital region with nothing between the facepiece cushion and
the face; <sup>i</sup>EMM, estimated marginal mean; <sup>j</sup>CI, confidence interval

The value of fit factor was significantly lower when the RPE was worn in the exercise state than when it was worn in the resting state. The fit factor was significantly the lowest among all wearing variations when RPE was applied by RPR_K. When fitted by RPR_R and RPR_H, the fit factor was significantly lower than when the PAPR was worn. No significant difference in fit factor values was observed when the PAPR was worn by PAPR_H, PAPR_K, and PAPR_R. There was no significant interaction between physical state and wearing variations.

## Discussion

In this study, we measured the fit factor of the RPR and PAPR used in the resting state and exercise state, using three different wearing variations. The fit factor of the RPR and PAPR in the exercise state was significantly lower than that during the resting state. This may be because the adhesion between the face and the RPE tends to decrease due to the exercising tasks. There was no significant interaction between the physical states and the wearing variations. It was suggested that even when workers wear RPE properly, working reduces respiratory protection. Since there is a possibility that working may impair protection, employers should try to reduce the risk by implementing engineering measures.

The fit factor of the PAPR used in the exercise state was greater than 100 in 9 of 10 cases for all wearing variations. There were three cases in which the fit factor of the RPR in the exercise state was greater than 100, using the recommended wearing method. There were no cases in which the fit factor of the RPR was greater than 100 when the knit cover was attached to the RPR. The inside of the PAPR was always under positive pressure, which may maintain respiratory protection even if working impairs the adhesion between the PAPR and the face. It is clear that PAPRs are superior to RPRs in terms of respiratory protection. However, it has been reported that PAPRs are rarely used in the workplace because^5^ they are expensive, they are larger than RPRs, and most workers complained about their heaviness;^11^ these reasons may have hindered the use of PAPR.

The mean fit factor of wearing a knit cover in RPRs was 12.3 in the resting state and 9.3 in the exercise state, which was significantly the lowest among all wearing methods. The fit factor was less than 100 in all participants, and this study also indicated that wearing a knit cover is a wearing method that can easily compromise the adhesion between the face and the RPR.^12^ When wearing a knit cover in PAPRs, the fit factor was more than 100 even during exercising tasks, and respiratory protection was maintained. If there is a risk of causing eczema or other skin problems by wearing RPE, workers are allowed to use a knit cover as long as there is good adhesion between the RPE and the face.^4^ Since this study showed that wearing a knit cover impairs adhesion to the RPR, it is recommended that workers with skin disorders who use a knit cover wear a PAPR.

There was no significant difference in the fit factor between the recommended method of wearing the RPR and the method of wearing the RPR over the helmet. In this study, we kept the pressure on the tightening cord constant among the participants, but there may be differences among individuals in the workplace. The RPR, PAPR, and helmet we examined were of one model and they were unused.

In this case, the RPE headband merely fits well with the helmet, and it is unclear whether the results are similar for the other models. It is also unclear how the degradation of the RPR or helmet affects the fit of the headband and helmet. Further study is required on these points, and we cannot yet recommend wearing RPE over a helmet. However, it has been reported that many workers in the workplace wore RPE over their helmets.^5^ Working in a dusty environment while wearing RPE with a poorly adherent attachment does not sufficiently prevent pneumoconiosis. It is important to provide continuous education and guidance to workers to ensure that they wear RPE with minimal leakage. Recently, the concept of workplace protection factor (WPF) has emerged because of the importance of evaluating the respiratory protection of RPE in the workplace environment. WPF is a measure of the protection provided in the workplace when a properly functioning respirator is correctly fitted and used,^13^ and several studies on WPF have been reported.^14-18^ In this study, respiratory protection during exercising tasks was evaluated in an artificial climate chamber, and it is desirable to evaluate the respiratory protection of various methods of wearing RPE in the workplace.

This study has some limitations. First, the number of participants was small, and type II errors may be large, so we need to increase the sample size. Second, the RPRs, PAPRs, knit cover, and helmet we used were of only one type, so we thought it was necessary to verify with multiple types of RPE. Moreover, the RPE, knit cover, and helmet used were new. In the workplace, workers use the equipment for many years, so the deterioration of the silicone of the RPE, knit cover, and helmet may affect the fit factor. Third, participants may have worn the RPE more rigorously than the actual workers. To minimize the effect of the individual’s ability to accurately fit the RPE, the participants wore the RPE while looking at a mirror reflection during the survey. If the examiner noticed an abnormality, such as a twisted strap, the participant was asked to remove the RPE and re-attach it. The verification of WPF in actual workers without any advice on how to wear the RPE is a subject for future study.

## Conclusion

We evaluated the respiratory protection of RPRs and PAPRs during exercising tasks, using various wearing methods. The fit factor of RPRs in the exercise state was significantly lower than that in the resting state, indicating inadequate respiratory protection. In contrast, the fit factor of PAPRs was significantly lower than that at rest, but respiratory protection was maintained. As in a previous study, PAPRs were found to be superior to RPRs in terms of respiratory protection. Therefore, PAPRs are better than RPRs for workers who have to wear RPE inappropriately due to health problems.

## Data Availability

Data are not available due to ethical restrictions.

## Acknowledgements

We would like to thank Editage (http://www.editage.com) for editing and reviewing this manuscript for English language.

## Declaration of Conflict interests

The authors declare no conflicts of interest.

## Author Contributions

Conceptualization: Taiki Shirasaka, Hajime Ando, Kazunori Ikegami

Data curation: Taiki Shirasaka, Hajime Ando, Kazunori Ikegami

Formal analysis: Taiki Shirasaka, Hajime Ando, Kazunori Ikegami

Funding acquisition: Akira Ogami

Investigation: Taiki Shirasaka, Hajime Ando, Kazunori Ikegami

Methodology – Development or design of methodology; creation of models.

Project administration: Taiki Shirasaka, Hajime Ando, Kazunori Ikegami

Supervision: Akira Ogami

Validation: Taiki Shirasaka, Hajime Ando, Kazunori Ikegami

Visualization: Hiroka Baba, Taiki Shirasaka, Hajime Ando, Kazunori Ikegami

Writing - original draft: Hiroka Baba, Hajime Ando, Kazunori Ikegami

Writing - review & editing: Hiroka Baba, Hajime Ando Kazunori Ikegami, Shingo Sekoguchi, Taiki Shirasaka, Akira Ogami

## Notes

**Funding** This study was funded by the Industrial Disease Clinical Research Grants.

### Competing Interest Statement

The authors have declared no competing interest.

### Funding Statement

This study was funded by the Industrial Disease Clinical Research Grants.

